# Genetic correlates of socio-economic status influence the pattern of shared heritability across mental health traits

**DOI:** 10.1101/2020.02.26.20028092

**Authors:** Andries T. Marees, Dirk J.A. Smit, Abdel Abdellaoui, Michel G. Nivard, Wim van den Brink, Damiaan Denys, Titus Galama, Karin J.H. Verweij, Eske M. Derks

## Abstract

Epidemiological studies show high comorbidity between different mental health problems, indicating that individuals with a diagnosis of one disorder are more likely to develop other mental health problems. Genetic studies reveal substantial sharing of genetic risk factors across mental health traits. However, mental health is genetically correlated with socio-economic status (SES) and it is therefore important to investigate and disentangle the genetic relationship between mental health and SES. We used summary statistics from large genome-wide association studies (average N∼160,000) to estimate the genetic overlap across nine psychiatric disorders and seven substance use traits and explored the genetic influence of three different indicators of SES. Using Genomic SEM, we show significant changes in patterns of genetic correlations after partialling out SES-associated genetic variation. Our approach allows the separation of disease-specific genetic variation and genetic variation shared with SES, thereby improving our understanding of the genetic architecture of mental health.

## Introduction

Substance abuse and psychiatric disorders pose major burdens on patients’ personal lives and that of their families, as well as on society as a whole. Previous studies reported robust evidence of high comorbidity between different mental health disorders, indicating that individuals with a diagnosis of one disorder are more likely to develop other mental health problems. Genetic studies may provide useful information on the contribution of shared genetic risk factors to the observed comorbidities, and thus potentially gives insights into shared underlying biology and pathology^1,2^. However, possible confounders such as socio-economic status (SES) may be correlated with genetic variation^3^ and should therefore be appropriately accounted for. Here, we assess the level of genetic overlap across 16 mental health traits, and explore the influence of SES-associated genetic variation on the pattern of shared heritability.

Twin and family studies have shown that genetic risk factors contribute substantially to the risk of developing a mental health disorder, with heritability estimates ranging between ∼40-80%^4-6^. Twin studies have further revealed significant genetic correlations across disorders, implying that partly overlapping genetic influences underlie vulnerability to different mental health traits (e.g.^7-10^). To better understand the biological basis of mental health disorders and their comorbidity, it is important to identify the specific genes that underlie these disorders. Genome-Wide Association Studies (GWAS) have become the standard approach to detect common genetic risk factors associated with psychiatric and substance use disorders and have been successfully applied to identify genetic loci for a wide variety of traits, including schizophrenia^11^, depression^12,13^, and lifetime cannabis use^14^. Historically, the primary aim of GWAS has been to identify genetic variants (single nucleotide polymorphisms; SNPs) that are statistically associated with complex traits such as human behaviour and disease risk^15^. More recent methodological innovations have enabled researchers to use genome-wide SNP effect sizes from GWAS to estimate the total proportion of phenotypic variation explained by all measured common genetic loci (SNP-based h^2^) and to assess the amount of genetic overlap across disorders^16,17^. The Brainstorm Consortium used GWAS data to estimate genetic correlations (r_g_) across ten psychiatric disorders, revealing considerable sharing of common genetic risk^1^. Genetic correlations were especially profound between schizophrenia and bipolar disorder (r_g_=.68), between major depression (MD) and anxiety disorder (r^g^=.79), and between obsessive-compulsive disorder (OCD) and anorexia nervosa (r_g_=.52). Vink et al. have extended this analysis to substance use phenotypes and showed large genetic overlap across psychiatric disorders and substance use phenotypes^18^. More recently, a study by the Psychiatric Genomics Consortium^2^ using GWAS summary statistics from eight psychiatric disorders, showed that the psychiatric disorders genetically cluster in three correlated factors: the first consisted primarily of disorders characterized by compulsive behaviours (anorexia nervosa, OCD, and Tourette Syndrome), the second by mood and psychotic disorders (major depression, bipolar disorder and schizophrenia), and the third by three early-onset neurodevelopmental disorders (autism spectrum disorder, ADHD, and Tourette Syndrome) as well as major depression. Unfortunately, substance use phenotypes were not included in this study.

However, a limitation of GWAS studies is that these are generally conducted within heterogeneous populations. Therefore, confounding factors, such as differences in environments, may bias genotype-phenotype associations and may therefore have an impact on estimates of SNP-based h^2^ and genetic correlations. For example, recent studies have shown that genetic associations may be inflated by population phenomena, such as population stratification^3,19^, gene-environment correlation^19,20^, and assortative mating^20^. Abdellaoui et al. showed significant geographic clustering of complex traits, even after controlling for ancestry^3^. They hypothesized that recent migration driven by educational attainment is one of the main contributing factors for this clustering of complex traits. One of the consequences of this geographic clustering is that alleles that are associated with educational attainment are correlated with environmental influences on health outcomes. This could cause an inflation of genetic correlation estimates between traits. To explore the potential influence of confounding factors at the family-level, Selzam et al. compared between-family and within-family genetic effects when using genome-wide polygenic scores to predict height, body mass index (BMI), intelligence, educational attainment, and ADHD symptoms^19^. They found that estimates of genetic effects were significantly reduced within families compared to between families, suggesting that unaccounted confounding factors inflate the between-family results. Interestingly, after controlling for family SES, the inflation largely disappeared in the between-family design, suggesting that SES is a major source of bias. Similar effects likely cause polygenic scores for educational attainment to be twice as predictive in non-adopted children as in adopted children^21^. These results highlight that genotype-phenotype associations based on samples of unrelated individuals may be confounded by external variables, especially by those that reflect complex social phenomena such as educational level^20^ and SES.

Low SES has been associated with increased levels of substance use and increased susceptibility for psychiatric disorders^22,23^. SES is often considered an environmental variable, but previous studies have shown that SES indicators have a substantial heritable component^24^ and that SES is an important factor to consider in relation to brain structure and function^25^. Twin studies have shown that 52% of the phenotypic variability in educational attainment is explained by genetic factors and large-scale GWAS have recently demonstrated that common genetic variants explain 11%, 21%, and 15% of the phenotypic variance of household income, social deprivation (i.e., Townsend index), and educational attainment, respectively^24,26^. Moreover, substantial genetic correlations between SES, substance use traits, and various psychiatric disorders have been found, with directions of effects in line with findings of traditional phenotypic epidemiology^23,24^. Estimates of the Brainstorm Consortium showed that ADHD, anxiety disorders, MDD, and Tourette syndrome show negative genetic correlations with intelligence and years of education whereas anorexia nervosa, autism spectrum disorders, and obsessive compulsive disorders show positive correlations^1^. The largest GWAS of smoking behaviour to date reported genetic correlations between smoking phenotypes and educational attainment of up to |0.55|^27^. Marees et al. recently presented findings suggesting that the genetic relation between alcohol consumption measures and mental health traits is mediated by SES^28^. In this study, we found that a high frequency of alcohol consumption (genetically associated with high SES) showed a different pattern of genetic correlations with mental health traits than high quantity of alcohol consumption (genetically associated with low SES). For example, alcohol frequency showed a negative genetic correlation with depression while alcohol quantity was positively correlated with depression. We hypothesized that these differences were due to the fact that the two alcohol measures were correlated with SES in opposite directions but did not formally evaluate this premise. Current findings suggest a strong genetic relation between mental health and SES which complicates the biological interpretation of estimates of shared genetic overlap.

In this study, we will test whether and to what extent SES-associated genetic variance influences genetic correlations across a range of mental health phenotypes, including nine psychiatric disorders and seven substance use phenotypes. While conventional genetic correlation analyses, such as cross-trait linkage disequilibrium score regression analysis (LDSC; as was used by the Brainstorm Consortium) only estimate conventional bivariate genetic correlations, we will apply a recently developed multivariate method called genomic structural equation modelling (genomic SEM^29^), which enables estimation of the joint genetic architecture of multiple complex traits. Genomic SEM formally models the genetic covariance structure of complex traits using GWAS summary statistics and allows the comparison of alternative multivariate genetic architectures. Using Genomic SEM, we can identify and partial out SES-associated genetic variation and separate this from the genetic variation that is not shared with SES. Separating these two sources of genetic variation will be relevant from a clinical perspective as the genetic risk that is “unique” for a mental health disorder will provide insight into disease-specific mechanisms while the genetic risk “shared” with SES will provide information on the contribution of general risk factors in a population.

We will use Genomic SEM to investigate the impact of genetic SES variance on the SNP-based heritability of 16 indicators of mental health and on the pattern of genetic correlations across these traits. The specific aims of this study are to i) estimate genetic correlations between three different indicators of SES and a composite SES factor with mental health traits; ii) determine to what extent genetic variation associated with SES contributes to estimates of SNP-based heritability of the mental health traits; and iii) elucidate what proportion of the genetic correlations between the various mental health traits is due to genetic overlap with SES.

## Methods

### GWAS summary statistics

The current study used existing summary statistics for psychiatric disorders included in the recent Brainstorm Consortium report^1^, expanded with the GWAS for substance use traits and SES indicator variables. Detailed information about the GWAS summary statistics, sample sizes, and availability are provided in Supplementary Table 1. For psychiatric disorders we used the case-control GWAS summary statistics for attention deficit/hyperactivity disorder (ADHD)^30^, anxiety/depression (AnxD)^31^, major depression (MD)^13^, bipolar disorder (BIP)^32^, schizophrenia (SCZ)^33^, autism spectrum disorder (ASD)^34^, obsessive-compulsive disorder (OCD)^35^, anorexia nervosa (AN)^36^, and Tourette’s syndrome (TS)^37^. In contrast to the Brainstorm Consortium report we excluded post-traumatic stress disorder, since this GWAS lacked power for our analyses.

For substance use traits, we used GWAS results from cannabis lifetime use (Cannabis)^14^, alcohol consumption frequency (AlcFreq)^28^, alcohol consumption quantity in subjects who drink at least once or twice a week (AlcQuan)^28^, ever initiated smoking (SmkInit)^27^, age of smoking initiation in ever-smoked subjects only (SmkAge)^27^, cigarettes per day (CigDay)^27^, and successful smoking cessation in lifetime smokers only (SmkCes)^27^. Note that SmkCes was coded such that 1 indicates a person has not quit smoking, whereas 0 indicates that he or she quit smoking.

For SES indicator variables we selected the GWAS for educational attainment (EA)^38^, household income (HI)^24^, and Townsend index (TI)^24^. All summary statistics are available for download, freely or by request/application.

### Estimates of genetic variance and genetic correlations

The proportion of trait variance explained by common SNPs (SNP-based heritability) for all SES, psychiatric and substance use traits was estimated using univariate LDSC^16^. We compared genetic variance estimates of the 16 mental health traits from a model excluding SES (i.e. SNP-based heritability) with those obtained in a model in which genetic SES variance was partialled out. SES was defined as a latent SES factor composed of educational attainment, household income, or Townsend index, but we also explored the influence of these SES indicator traits in isolation.

Genetic correlations between pairs of traits were estimated using bivariate LDSC^17^. LDSC is robust against confounding due to population stratification, and against full or partial sample overlap and cryptic relatedness across GWAS samples. LDSCs in the current study were performed using 1,215,002 SNPs present in the HapMap 3 reference panel, with exclusion of the major histocompatibility complex (MHC) region on chromosome 6. In assessing whether a genetic correlation was significant, we used a Benjamini-Hochberg FDR correction to account for multiple testing: the 120 genetic correlations between all 16 mental health traits before partialling out genetic SES variance, and then for the 120 genetic correlations between all 16 mental health traits after partialling out the genetic SES variance, (i.e., applying the FDR outcome in five different sets of 120 *p*-values).

### Genetic modelling

To estimate SNP-based heritabilities and genetic correlations with the effects of the latent SES factor partialled out, we used Genomic SEM^29^ R package v0.0.2 (https://github.com/MichelNivard/GenomicSEM/wiki). The SEM models are shown in Figure 1. Each Genomic SEM model includes five traits: the two mental health traits and the latent SES factor which is represented by three SES indicator traits. We fit models for all possible combinations of the 16 mental health traits. Genomic SEM produces genetic covariance matrices using a multivariable extension of LDSC for all of the input variables in a model - in the current model a 5×5 matrix - with SNP-based heritability on the diagonal and genetic correlations off-diagonal. This multivariable genetic covariance matrix is entered into the structural equation modelling R-package lavaan 0.6-3 (Rosseel, 2012; doi: 10.18637/jss.v048.i02). The SES residual model fitted a single latent common factor that represents the overlapping genetic variance between the three SES indicator traits, which has the advantage of excluding variable-specific variance (e.g., variance present in educational attainment but not present in the other SES indicator traits). The latent SES factor variance is then used to regress out variance from the observed genetic variance of the two mental health traits of interest. In other words, the model removes the effect of genetic SES variance on both SNP-based heritability of each of the two target traits and their genetic covariance. The three SES indicator traits were also tested separately for their effect on the pairwise genetic correlations in a simplified 3×3 model that did not construct a latent SES factor, but directly regressed the effect of a SES indicator traits from the two mental health traits in the model.

**Figure 1.**
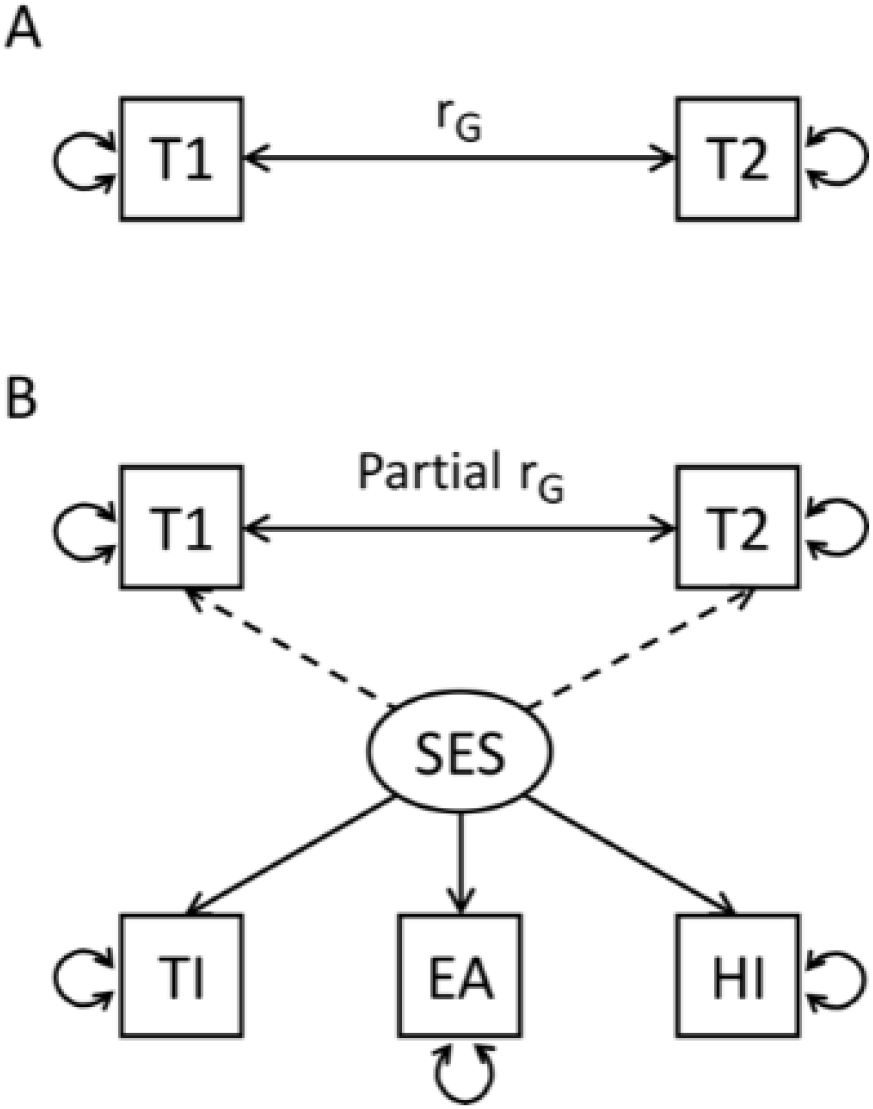
Genomic SEM path model fitted on the genetic covariation matrices estimates the genetic correlation (solid bidirectional arrow) between two traits of interest 1 and 2 (T1, T2). (A) path model of the simple bivariate genetic correlation using LDSC. (B) The model including a latent SES factor which represents the shared genetic variation of SES indicator traits Townsend Index (TI), Educational Attainment (EA), and Household Income (HI) (solid directional arrows). The latent SES factors’ genetic variance is regressed out from the variance of T1 and T2 (dashed arrows) affecting the estimates for heritability and genetic correlation.

### Significance of SES-induced change in genetic correlations

To assess the significance of the effect of removing SES-associated genetic variance on the genetic overlap of pairs of traits, we used a Monte Carlo random sampling technique based on the model estimates and their variability. In Genomic SEM, variability of the estimates is obtained by jack-knifing each chromosome (leave-one-chromosome-out) and re-estimating all free parameters in the model. Each jack-knife iteration results in a full symmetric (5×5) matrix with 15 free parameters representing the SNP-based heritabilities and genetic correlations of the two traits of interest and the three SES indicator traits. Based on these jack-knifed estimates, Genomic SEM provides a standard error (SE) for the 15 parameter estimates plus a term for the covariation between the estimates, resulting in a (15×15) matrix that captures not only parameter error but also error covariation, which tends to be substantial. Disregarding such correlated error would lead to substantially increased type-II errors. Using the matrix of estimates and the matrix of errors and error-covariances, we created 1000 samples using the mvrnorm function (MASS package in R^39^) from multivariate normal distributions of the estimates which followed the error-covariance structure. Each sampled matrix was used to recalculate SNP-based heritability and genetic correlation in the model with and without the SES factor. Significance was established by noting the change in estimates of genetic correlation, comparing estimates before and after SES factor inclusion across the 1000 models computed from the sampled data. We counted the number of times an estimate changed in the opposite direction as in the observed models (for example, if an r_G_ was lower after including SES in the full, unsampled model, we counted the number of occasions in the 1000 sampled models that the inclusion caused an increase in r_G_). P-values were defined as this count divided by the 1000 samples, multiplied by two for two-sided testing.

The process was repeated for each of the SES indicator traits separately. Benjamini-Hochberg FDR was used to correct for multiple testing for the 120 genetic correlations between all 16 mental health traits after partialling out the SES-factor (i.e., applying the FDR outcome in five different sets of 120 *p*-values).

### Graph analysis of genetic correlation matrices

To establish the effect of SES on genetic clustering across substance use and psychiatric phenotypes, we used graph analysis on the squared genetic correlation matrices before and after removing the SES-associated genetic variance. R package iGraph (v1.2.4.1)^40^ was used to visualise the connectivity strength based on the proportion of variance explained (i.e. squared genetic correlation). Clusters were predefined as substance use traits (AlcFreq, AlcQuan, Cannabis, CigDay, SmkCes, SmkInit, AgeSmk) and psychiatric traits (ADHD, AN, AnxD, ASD, BIP, MD, OCD, SCZ, TS). The effect of removing SES genetic variance was established for eigenvector centrality measure for each vertex. The effect on clustering was established by comparing intra- and inter-cluster connection strength (r-squared) for the predefined clusters (psychiatric and substance use). Finally, we assessed whether genetic clustering was increased using the fast greedy algorithm of Newman and Girvan^41^ without predefined clusters.

## Results

We found substantial genetic correlations between the three SES indicator variables: educational attainment-household income r_g_=0.86, SE=0.04; educational attainment-Townsend Index r_g_=0.49, SE=0.03; household income – Townsend Index r_g_=0.77, SE=0.05. The SNP-based heritability estimates for the latent SES factor, educational attainment, household income, and Townsend Index were 0.06 (SE=0.005), 0.09 (SE=0.002), 0.06 (SE=0.003), and 0.03 (SE=0.002), respectively. Also see Supplementary Tables 5 and 6.

### Genetic correlations of SES with the mental health traits

Figure 2 and Supplementary Table 1 show the genetic correlations between the latent SES factor with the 9 psychiatric disorders and 7 substance use traits. All 16 traits showed significant genetic correlations with the latent SES factor, with negative correlations (N=9) being slightly more common than positive correlations (N=7). Substantial negative genetic correlations (r_g_< - 0.4) of the latent SES factor were found with ADHD, anxiety disorder, major depression, smoking initiation, cigarettes smoked per day, and smoking cessation, whereas substantial positive genetic associations (r_g_> 0.4) were found with frequency of alcohol consumption and age at smoking initiation (i.e. genetic variants underlying younger age of smoking were associated with genetic variants for lower SES).

**Figure 2:**
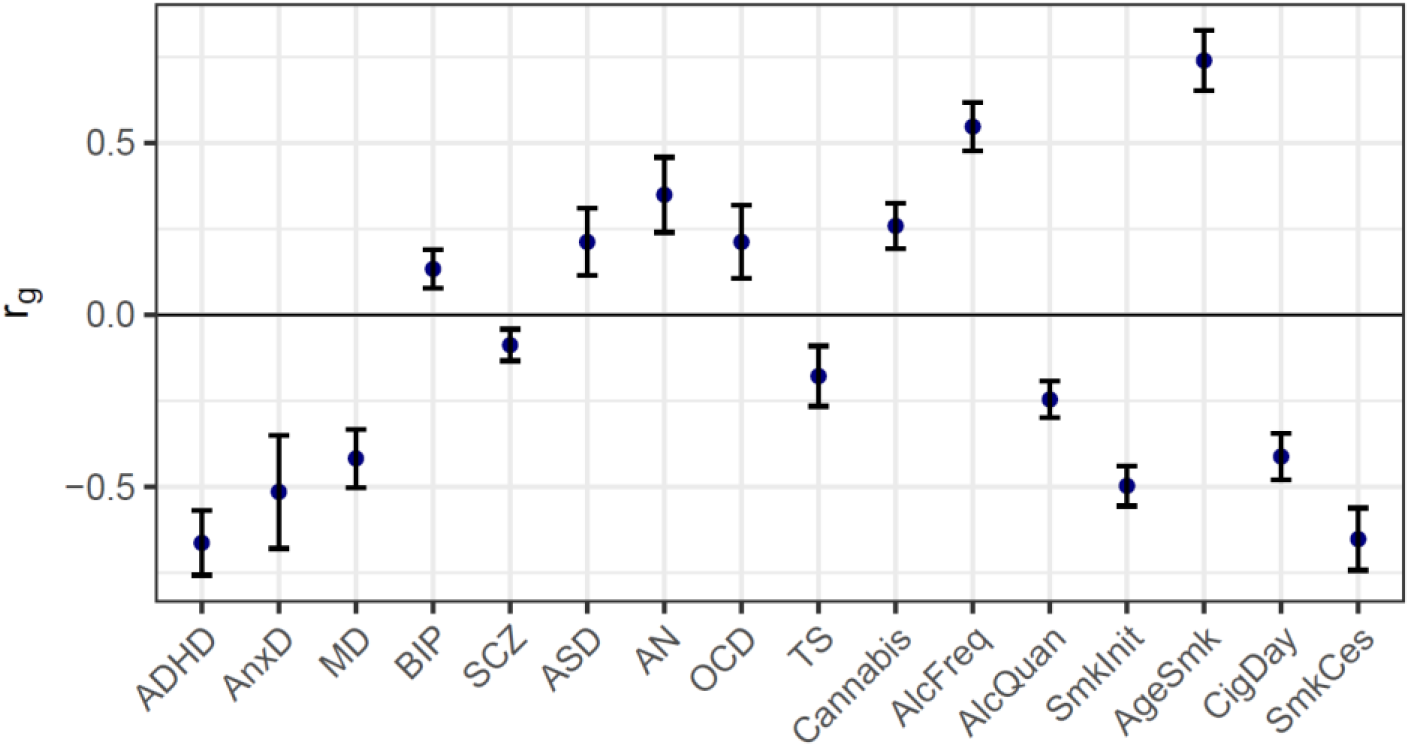
Genetic correlations between the latent SES factor and 9 psychiatric disorders and 7 substance use traits as computed with LDSC (error bars show ± 2×SE).

Genetic correlations were also estimated with each of the individual SES indicator traits (i.e., household income, Townsend index, and educational attainment); results were very similar to those for the latent SES factor (Supplementary Table 2 and Supplementary Figure 1).

### Attenuation of SNP-based heritability of mental health trait through SES

Figure 3 and Supplementary Table 3 show the SNP-based heritability estimates of the 16 mental health traits as well as the residual genetic variance after partialling out genetic effects that are shared with the latent SES factor. A reduction in genetic variance was most apparent for ADHD with attenuation of 43% after removing genetic SES variance. Genetic variance was also reduced for anxiety disorder, major depression, anorexia nervosa, frequency of alcohol consumption, smoking initiation, age at smoking initiation, and smoking cessation. The smallest reductions were observed for e.g. bipolar disorder, schizophrenia, and quantity of alcohol use.

**Figure 3:**
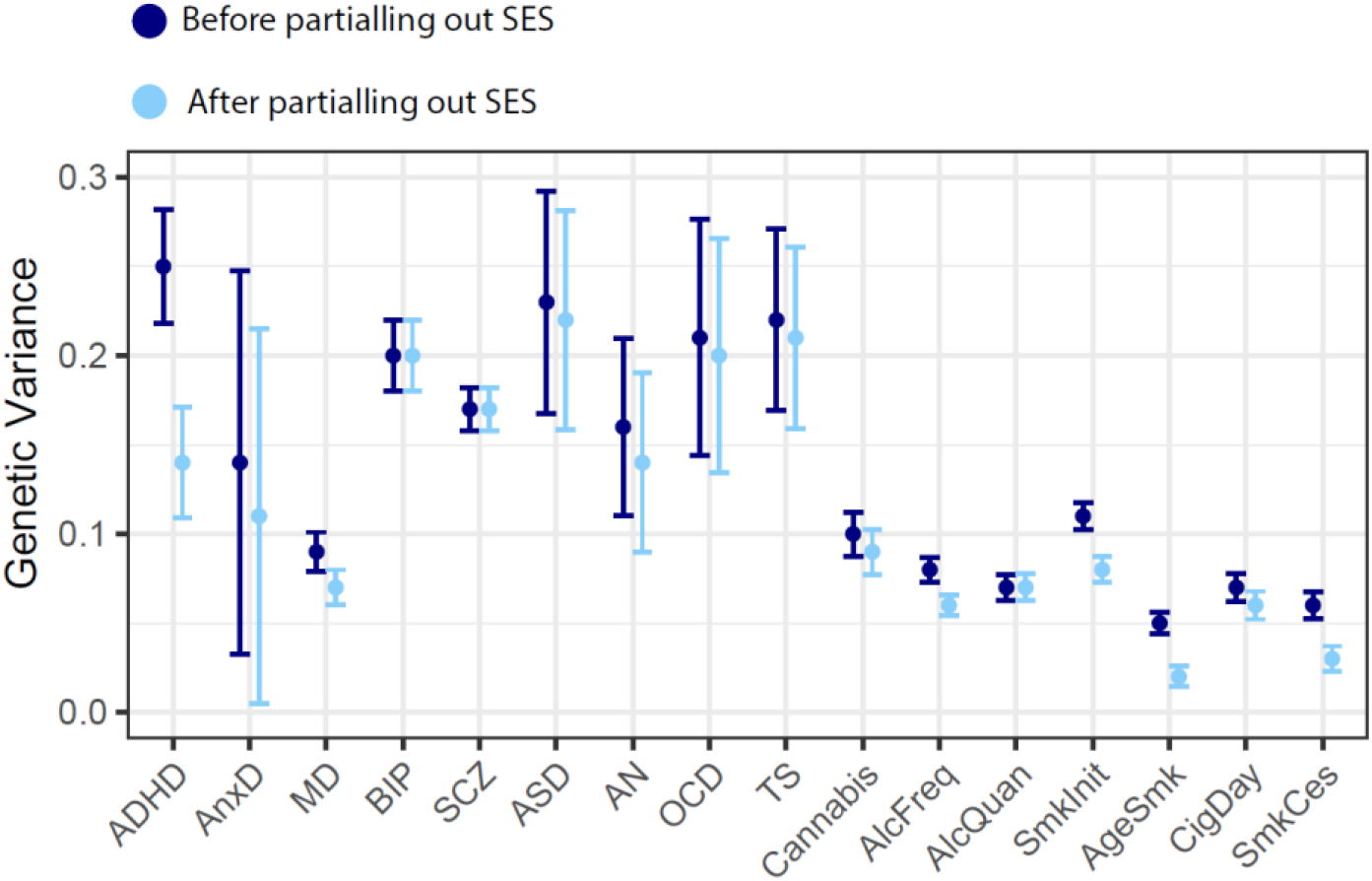
Genetic variance explained by SNPs before (SNP-based heritability) and after removing genetic effects overlapping with the latent SES factor (error bars show ± 2×SE).

Results were roughly similar when examining the effect of individual SES indicator traits on the SNP-based mental health trait heritabilities, although the attenuation of the heritability estimates was generally smaller (Supplementary Table 3 and Supplementary Figure 2).

### Proportion of the genetic correlations between the mental health traits before and after removing SES-associated genetic variance

Figure 4 and Supplementary Figure 3 show genetic correlations between the mental health traits before and after removing genetic variance in common with the latent SES factor. The direction of change in genetic correlation is dependent on the direction of the genetic correlation between SES and the two traits: if both traits were genetically correlated with SES in the same direction (i.e. both positive or both negative), the genetic correlation between the traits decreased when partialling out the latent SES factor, whereas if the two traits showed genetic correlations with SES in opposite directions, the genetic correlation between the two traits increased. Exact estimates of the genetic correlations before and after partialling out the effects of SES can be found in Figure 4, and Supplementary Table 4.

**Figure 4:**
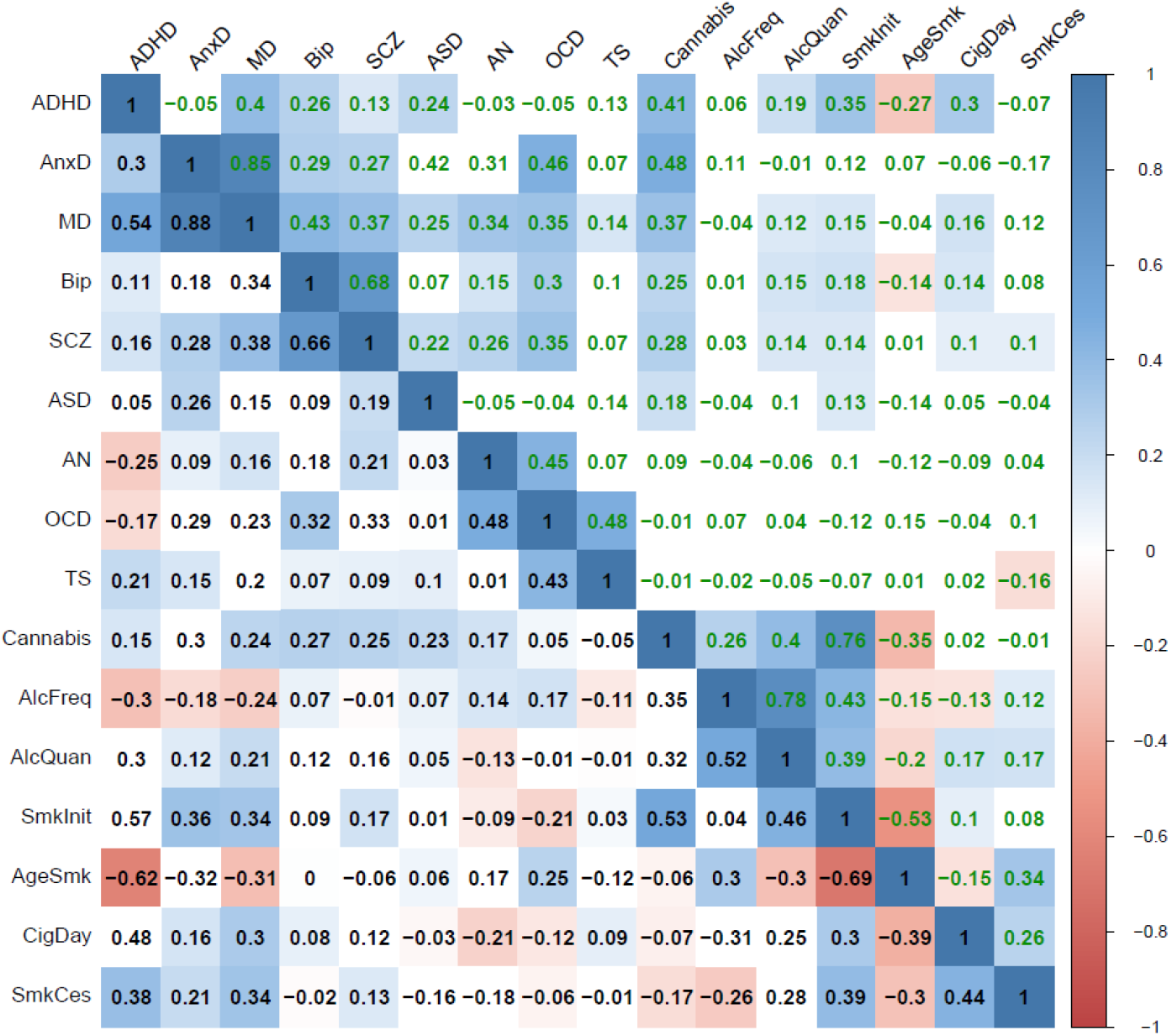
The genetic correlations before (lower diagonal, in black font) and after (upper diagonal, in green font) partialling out latent genetic SES factor variance. Coloured squares indicate significant genetic correlations (FDR corrected).

Notably, genetic correlations were most strongly altered for ADHD, consistent with the strong reduction in SNP-based heritability of ADHD in Figure 3. Genetic correlations with (a subset of) other traits also changed considerably for anxiety disorder, major depression, frequency of alcohol consumption, age at smoking initiation, and smoking cessation. Changes in the pattern of genetic correlations were negligible, on the other hand, for bipolar disorder, schizophrenia, autism spectrum disorder, obsessive-compulsive disorder, and Tourette syndrome (although high statistical power that comes with some of the summary statistics resulted in statistically significant changes even when the changes in point estimates were minimal).

When examining the results of the effects of the individual SES indicator traits on the genetic correlations (Supplementary Figures 4-10, and Supplementary Table 4), the changes in genetic correlations between trait pairs are very similar for the different SES indicator traits. However, many more changes are significant after partialling out educational attainment than when removing the effects of household income, Townsend Index, or the latent SES factor.

### Effect of SES on genetic clustering

Figure 5 shows the genetic correlation clusters based on graph analyses of genetic variance explained before and after removing SES-associated genetic variation. The graphs indicated evidence for stronger clustering of the psychiatric disorders after SES genetic variance removal, that is, comparatively to substance use traits. This was caused by strongly decreased edge weights (in terms of variance explained) between substance use traits (–0.062), a marginal decrease in edge weights between psychiatric traits (–0.010), and decreased edge weights across substance use and psychiatric traits (–0.023). These changes resulted in a clearer separation of the substance use and psychiatric cluster and stronger cohesion within the psychiatric cluster post-SES-removal (Figure 5 right). Using the Newman and Girvan (2004) clustering algorithm^41^, we observed and increased modularity Q (from 0.251 to 0.321). This algorithm separated one substance use cluster and two psychiatric clusters but note that ADHD is a notable exception and clustered to substance use traits, even after removing genetic SES variance. The two psychiatric clusters changed from (autism spectrum disorders, anxiety, major depression) and (anorexia nervosa, OCD, Tourette’s syndrome, schizophrenia, bipolar disorder) to (autism spectrum disorders, anxiety, major depression, anorexia nervosa, OCD, Tourette’s syndrome) and (schizophrenia, bipolar disorder).

**Figure 5.**
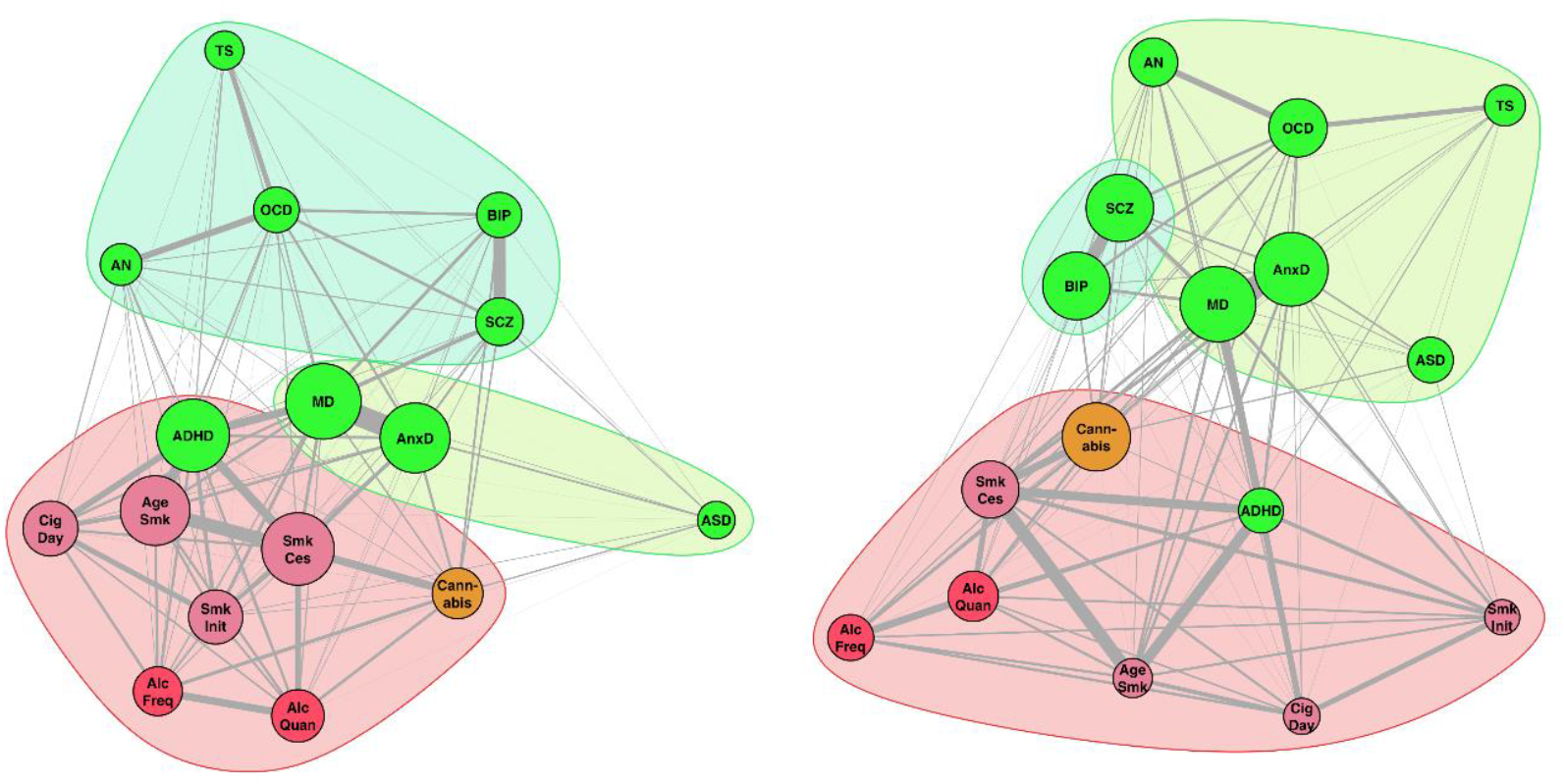
SNP-based genetic correlations between the mental health traits illustrated with a graph to reveal complex genetic relationships. Each vertex (node) represents a mental health trait, with green nodes representing psychiatric disorders and reddish nodes the substance use traits (red for alcohol, orange for cannabis, and lilac for smoking related traits). Vertex size is based on eigenvector centrality with a minimum offset. Weighted undirected graphs were created with the absolute genetic correlation as connection strength (represented in line thickness). Vertex layout was based on the Fruchterman and Reingold algorithm42. **Left;** Graph before removing genetic SES variance resulted in three clusters (Clustering coefficient Q=0.251). **Right;** Graph after removing genetic SES variance showed an increased clustering index (Q=0.321), reshaped psychiatric genetic clusters, and kept the substance use cluster intact (including ADHD).

## Discussion

We have used the summary statistics from large-scale GWASs (average N ∼160,000) to examine the extent to which genetic overlap with SES influences genetic variance in and genetic overlap across 16 mental health phenotypes. We show that removing the variance of the latent SES factor significantly changes the pattern of genetic relationships between mental health traits.

### Genetic overlap with latent SES factor

All 16 mental health traits showed a significant genetic correlation with a latent SES factor extending findings of previous studies^1,14,18,27^. Although most of the 16 genetic correlations between mental health traits and the latent SES factor were negative, in seven of the 16 genetic correlations the genetic propensity for lower SES was associated with an decreased genetic risk for psychiatric traits, namely for OCD, bipolar disorder, autism, anorexia nervosa, alcohol intake frequency, and cannabis use. Some of these positive genetic correlations are in line the phenotypic correlations reported by epidemiological studies, such as the positive phenotypic correlations of SES with autism^43^ and anorexia nervosa^44^; studies have been less consistent about the direction of the relationship between bipolar disorder and OCD with SES^45^. The potential mechanisms behind these findings can be diverse, for example, the positive genetic correlation between educational attainment and cannabis use (rG= .36) could potentially be related to higher rates of cannabis use in metropolitan vs. rural areas^46^ supporting the previously reported association between lifetime cannabis use and higher childhood family SES^47^.

### Genetic overlap between mental health traits

The significant genetic correlations between mental health and SES and the general reduction of genetic cross-trait correlations after removing genetic SES variance suggest that part of the heritability of mental health as well as the genetic overlap between the different mental health traits is due to shared genetic variation with SES. For pairs of traits with opposite directions in their genetic correlation with SES, an increase in genetic correlation was observed. For example, the genetic correlation between ADHD (r_g_ with SES = -0.64) and lifetime cannabis use (r_g_ with SES = 0.25) was 0.15 and increased to 0.31 after partialling out SES genetic variance. This shows that there are instances in which genetic overlap between traits is obscured by their shared genetic overlap with SES, highlighting the complex interdependence among these variables.

The relevance of our approach lies in the ability to compare patterns of genetic correlation before and after removing SES-associated genetic variation. For example, we previously reported that frequency and quantity of alcohol consumption are genetically correlated with SES in opposite directions and hypothesized that this may explain the different patterns of genetic correlations between the two alcohol measures and mental health traits^28^. Indeed, after removing genetic variance associated with SES, the genetic correlation between these two alcohol measures increased while the pattern of genetic correlations with other traits became much more similar. In the bivariate analyses that do not include the influence of SES, frequency of alcohol consumption was genetically associated with lower risk of ADHD, anxiety, and depression. However, after partialling out SES-associated genetic variation, the genetic correlations between frequency of alcohol consumption and these aspects of mental health were negligible. This suggests that the mental health benefits of moderate drinking may reflect SES confounding rather than direct causal effects of drinking behaviour.

Using graph analyses to present clustering of the mental health traits, we found three genetic clusters. Interestingly, ADHD clustered within the substance use category, rather than in one of the psychiatric clusters. After partialling out the SES genetic variation, ADHD remained in the substance use cluster. This indicates that some neurobiological causes of ADHD are shared with substance use traits above and beyond SES genetic variance—which includes the possibility of direct causal links. The results showed that genetic overlap among mental health traits partly depends on the overlap with genetic SES variation. This has consequences for conceptualizations of an underlying psychopathology factor (i.e., *p*-factor).^2,29,48^

While the strength of genetic association within the substance use cluster decreased on average after partialling out SES, the average strength of genetic association within the two psychiatric disorder clusters did not change substantially. However, the effect of partialling out SES had strong effects on some genetic associations within the psychiatric clusters (either becoming stronger or weaker) but not on others (also see Figure 4). This resulted in a reordering of the psychiatric disorders clusters. For traits that were highly affected by SES, in particular MDD and ADHD, their contribution to a shared psychopathology factor (see e.g. ^2^) must be considered and interpretation may be different when spurious contributions, such as regional effects associated with SES^3^, are removed. Also, as some genetic associations between psychiatric disorders changed sign or increased in strength after partialling out SES genetic variance, our analyses show that confounding factors may obscure genetic overlap between traits.

### Mechanisms behind the effects of shared genetic SES variation

There are different mechanisms that can explain why SES-associated genetic variation influences GWAS findings of mental health traits and the observed patterns of genetic correlations. First, lower SES is geographically clustered^3^, and is thus related to a wide range of detrimental environmental variables that may increase the risk for both physical and mental health problems as well as substance use. Living in disadvantaged neighbourhoods may place individuals at risk for substance use through increased availability and targeted marketing for alcohol products^49,50^. Adverse neighbourhood circumstances are also associated with increased risk of developing other psychiatric disorders^51,52^. It has been shown recently that a genetic predisposition for higher educational attainment is strongly associated with migration to better neighbourhoods with fewer exposures to harmful environmental influences^3^.

Second, a causal influence of SES on mental health could also exist at a neurocognitive level, since lower cognitive abilities are correlated with lower educational attainment, lower income^53,54^, and lower impulse control^55^. The literature demonstrates that lower cognitive abilities and lower impulse control are correlated with both increased substance use and increased risk for psychiatric disorders^55-57^. Third, these findings may also partly reflect pleiotropy in which genetic variants influence the risk of mental health traits as well as SES through an underlying *p*-factor shared among substance use, psychiatric disorders, and lower SES^48^. Our current results provide information on the extent to which genetic correlations across mental health traits change after controlling for SES, but do not allow us to separate between these alternative mechanistic explanations.

### Educational Attainment

When considering the separate components used to derive the underlying SES factor, namely educational attainment, household income, and Townsend index, the strongest effects were observed for educational attainment. Nearly all genetic correlations were significantly altered after partialling out genetic variation associated with educational attainment (Supplementary Figure 7). This may be related to the strength of the signal of the educational attainment GWAS, which has a much larger sample size (N=766,345) than the other SES indicators (N=96,900 and N=112,005). However, it may also reflect the importance of educational attainment on life outcomes related to both SES and mental health^58^.

### Implications

SES is an important factor to consider when interpreting the results of GWAS studies and post-GWAS analyses. Results of GWAS that are uncontrolled for SES-associated genetic variation may lead to biased estimates of trait-specific variance. The range of phenotypes for which this applies, is likely more extensive than those covered in the present study, e.g., traits such as BMI, longevity, and cardiovascular diseases, may be influenced in a similar way. Depending on the specific purpose of a genetic study, researchers can consider to partial out SES-associated genetic variance thereby reducing SNP-based heritabilities for traits that are genetically linked to SES, but the remaining genetic variance will be more trait-specific, and possibly more relevant from a clinical perspective.

Removing SES related genetic variance in genetic studies will also influence results of secondary analyses that use the GWAS summary statistics, such as Polygenic Risk Score (PRS) analyses^15^. Partialling out SES may reduce the predictive power of PRS analyses as it removes part of the genetic variance, but the remaining genetic variance will more specifically reflect the phenotype of interest, which may increase their potential clinical utility.

Our results also suggest a possible violation of an important assumption of Mendelian Randomization (MR) analyses, which states that the genetic variants included in the instrumental variable should be independent of factors (measured and unmeasured) that confound the exposure-outcome relationship^20,59^. SES is likely to be a confounder of MR analyses through its genetic associations with the genetic instruments for mental health phenotypes, which may affect results of MR studies aimed at establishing causal relationships across mental health traits, and other complex traits (e.g., BMI, longevity, and cardiovascular diseases).

Our findings have possible implications for the discussion regarding the diagnostic boundaries across mental health disorders. It has previously been suggested that the observation of strong genetic correlations across psychiatric disorders suggests horizontal pleiotropy^1^, which would be indicative of a mismatch between current clinical boundaries and the underlying pathogenic processes. We show that the genetic comorbidity between mental health traits is more complex and that the genetic clustering of traits changes when partialling out genetic variance shared with SES.

### Limitations

Our findings need to be interpreted in the context of some limitations. First, SES is a multifactorial concept and there is no consensus in the field on how to best measure SES^25^. However, we used GWAS summary statistics to generate a latent SES factor composed of three indicators of SES, using both self-reported (educational attainment) and more objective measures (Townsend Index). We have further performed additional analyses to explore the influence of each of these individual indicators and found effects to be largely consistent. This provides an indication that our selection of SES indicators is not critical for the observed changes in genetic correlations and SNP-based heritability.

Second, as is the case for most genetic studies on mental health, the summary statistics in this study are based on samples with an overrepresentation of participants from Western, Educated, Industrialized, Rich, and Democratic (WEIRD) populations. This reduces the generalizability of our findings and future studies should be focused on exploring the relations between SES and mental health in other populations.

Finally, while Genomic SEM is much more flexible in including the effects of confounding factors in the Structural Equation Model, the nature of our data does not allow us to draw conclusions on the nature of the causal relationships between mental health phenotypes. While SES may confound the relations across mental health phenotypes, it is also possible that some of these phenotypes have a direct causal influence on SES (e.g., ADHD, MDD, and alcohol use may lead to lower SES due to either lower educational attainment or lower income). This will make SES a collider rather than a confounder. Correcting for SES as a collider will bias estimates of correlation between two traits, which could explain some of the shifts we reported. Whether SES is a collider, confounder, or shows bidirectional causality with the psychiatric traits may be investigated using genetic causal modelling.

### Conclusion

Our findings reveal that SNP-based heritabilities of 16 mental health traits, and the genetic correlations between them, are influenced by genetic overlap with SES traits. Our findings suggest that the genetic overlap between substance use traits and psychiatric disorders^1,14,27,60^ is in part due to their shared genetic overlap with SES. These findings provide important insights into the complexity of these associations and highlight the need to consider the role of SES in future studies investigating the genetic basis of mental health traits.

## Data Availability

The GWAS summary statistics used in this article are all publicly available (see references of the GWAS articles).

## Acknowledgements

A.A., A.T.M., & K.J.H.V. are supported by the Foundation Volksbond Rotterdam. A.T.M. & T.J.G. are supported by the Netherlands Organization for Research (NWO) Vidi grant 0.16.Vidi.185.044. M.G.N. is supported by ZonMw grants 849200011 and 531003014 from The Netherlands Organisation for Health Research and Development. This research was supported by the National Institute on Aging, under grants RF1055654 and R01AG058726.

## Supplementary Figures

**Supplementary Figure 1:**
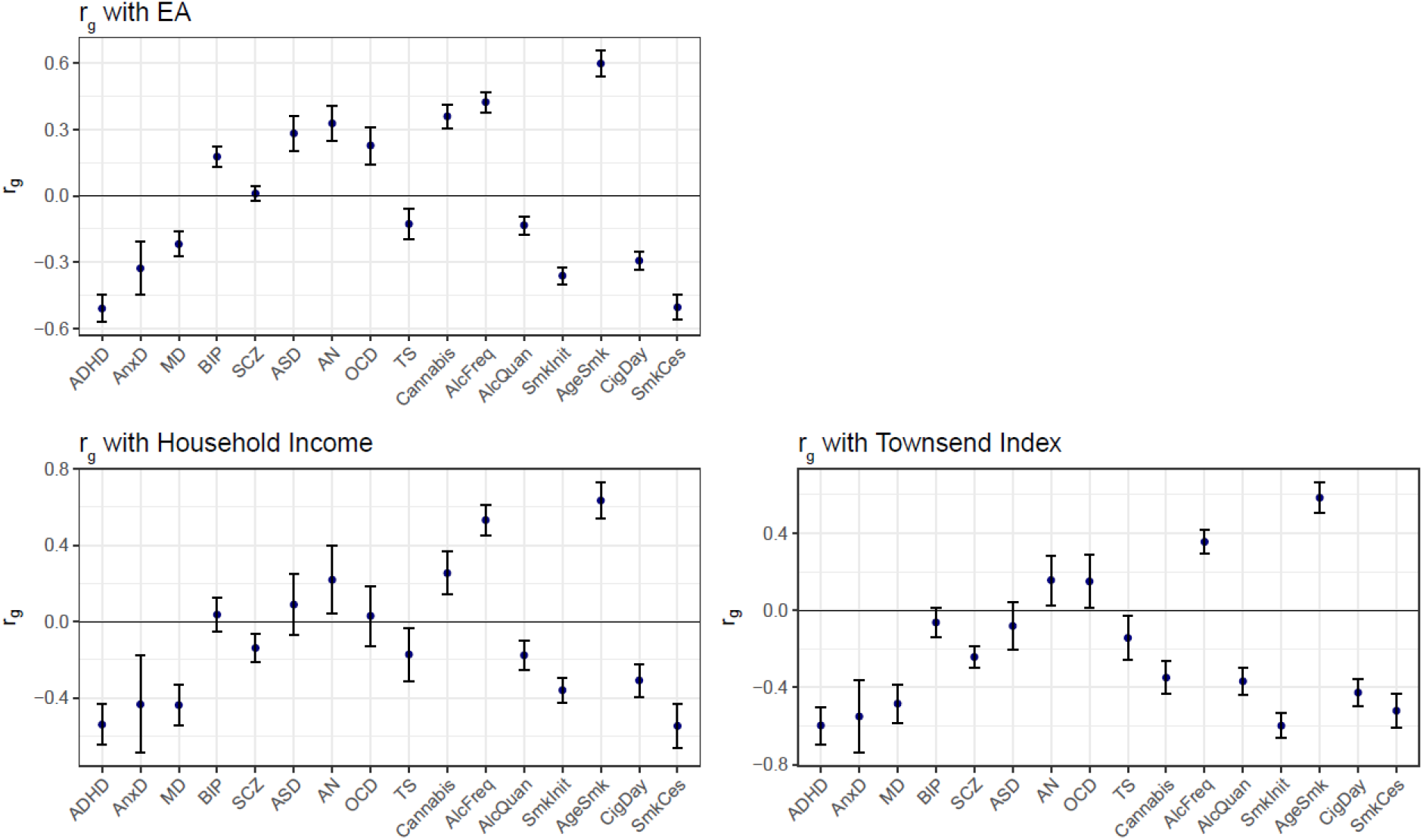
Genetic correlations between SES indicators and 9 psychiatric disorders and 7 substance use traits as computed with LDSC (error bars show ± 2×SE).

**Supplementary Figure 2:**
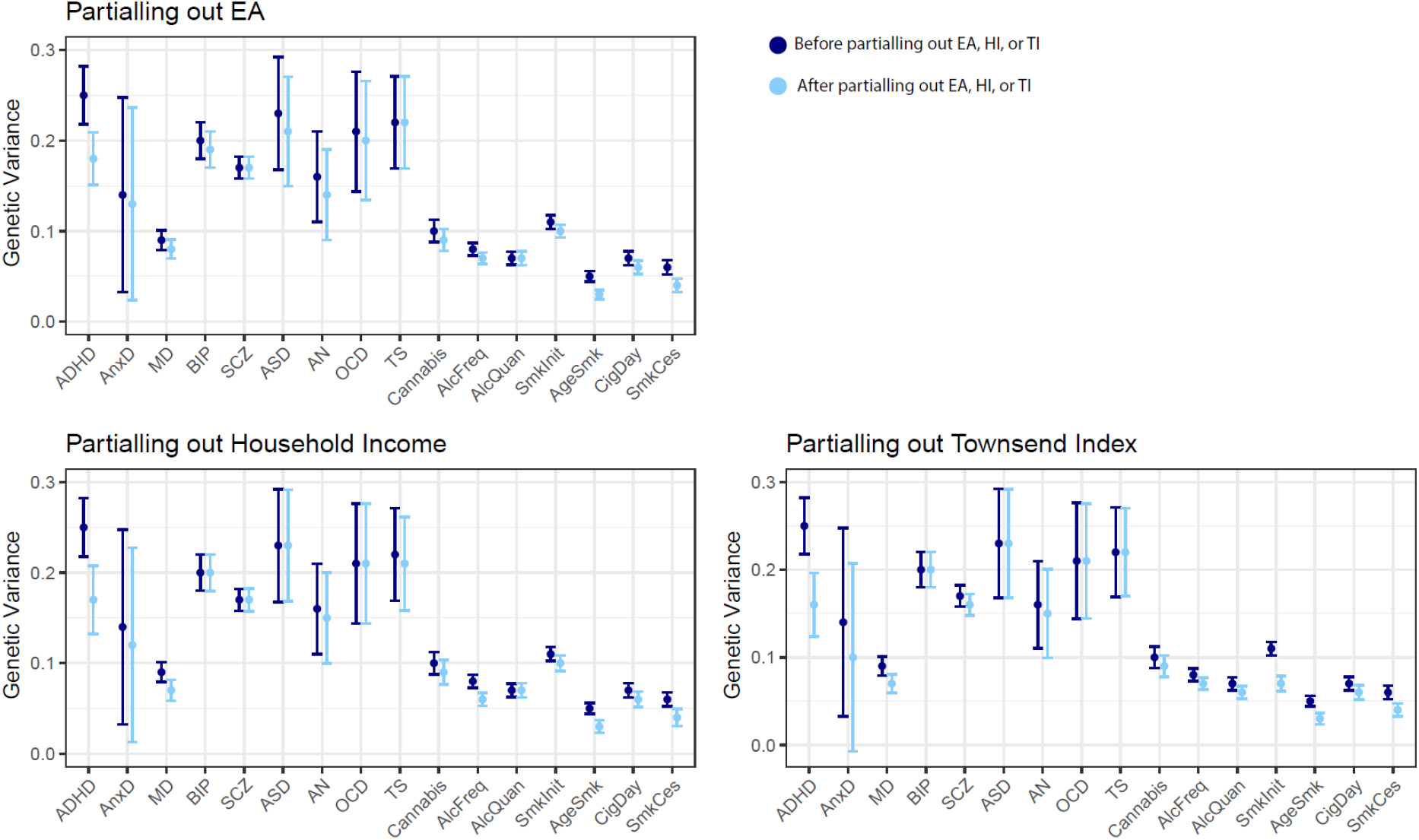
Genetic variance explained by SNPs before (SNP-based heritability) and after removing genetic effects overlapping with the SES indicators (error bars show ± 2×SE).

**Supplementary Figure 3:**
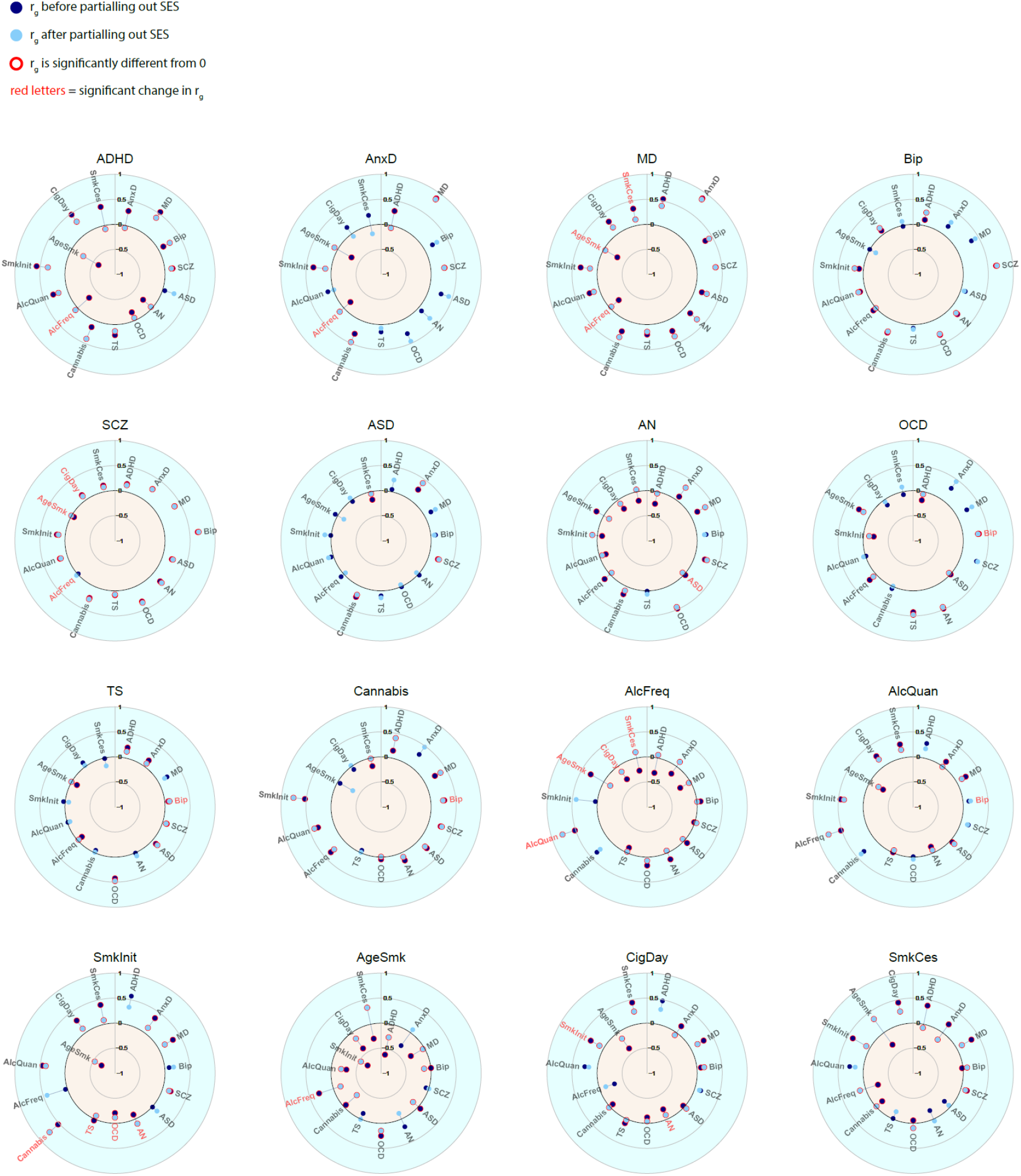
Genetic correlations before and after partialling out the SES factor. Significant genetic correlations are indicated with red circles and significant changes in genetic correlations after partialling out SES are indicated in red letters.

**Supplementary Figure 4:**
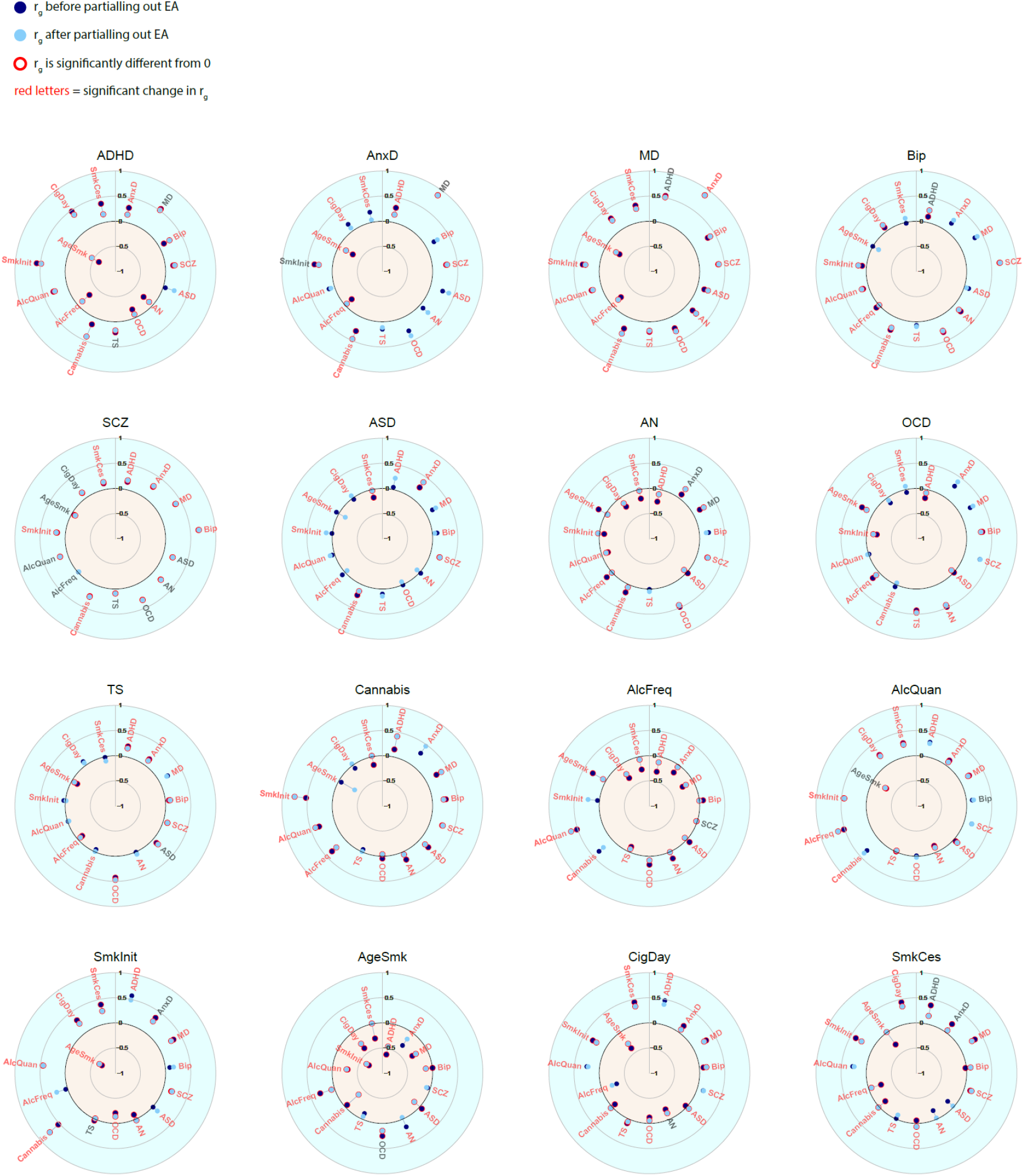
Genetic correlations before and after partialling out educational attainment (EA). Significant genetic correlations are indicated with red circles and significant changes in genetic correlations after partialling out EA are indicated in red letters.

**Supplementary Figure 5:**
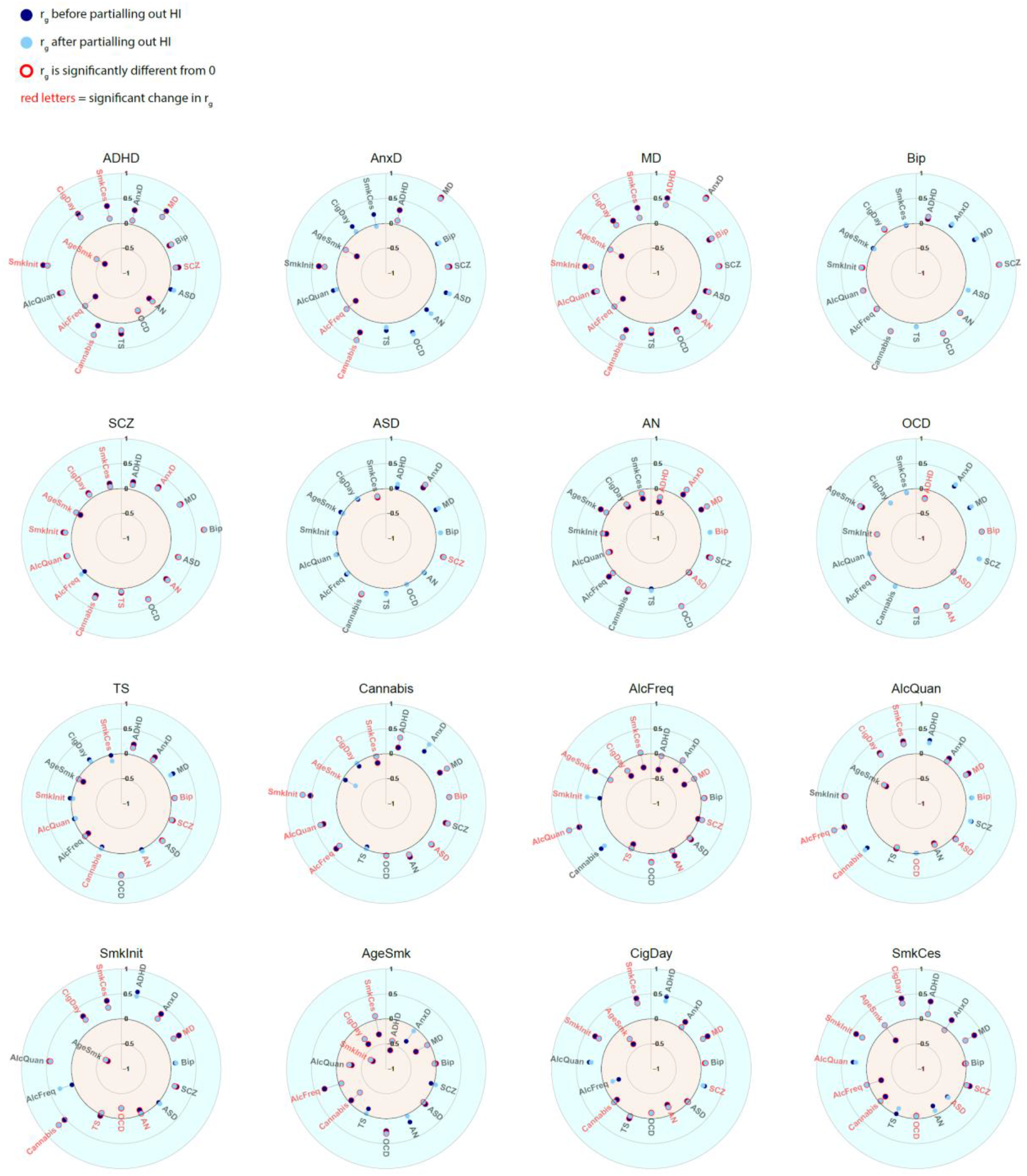
Genetic correlations before and after partialling out household income (HI). Significant genetic correlations are indicated with red circles and significant changes in genetic correlations after partialling out HI are indicated in red letters.

**Supplementary Figure 6:**
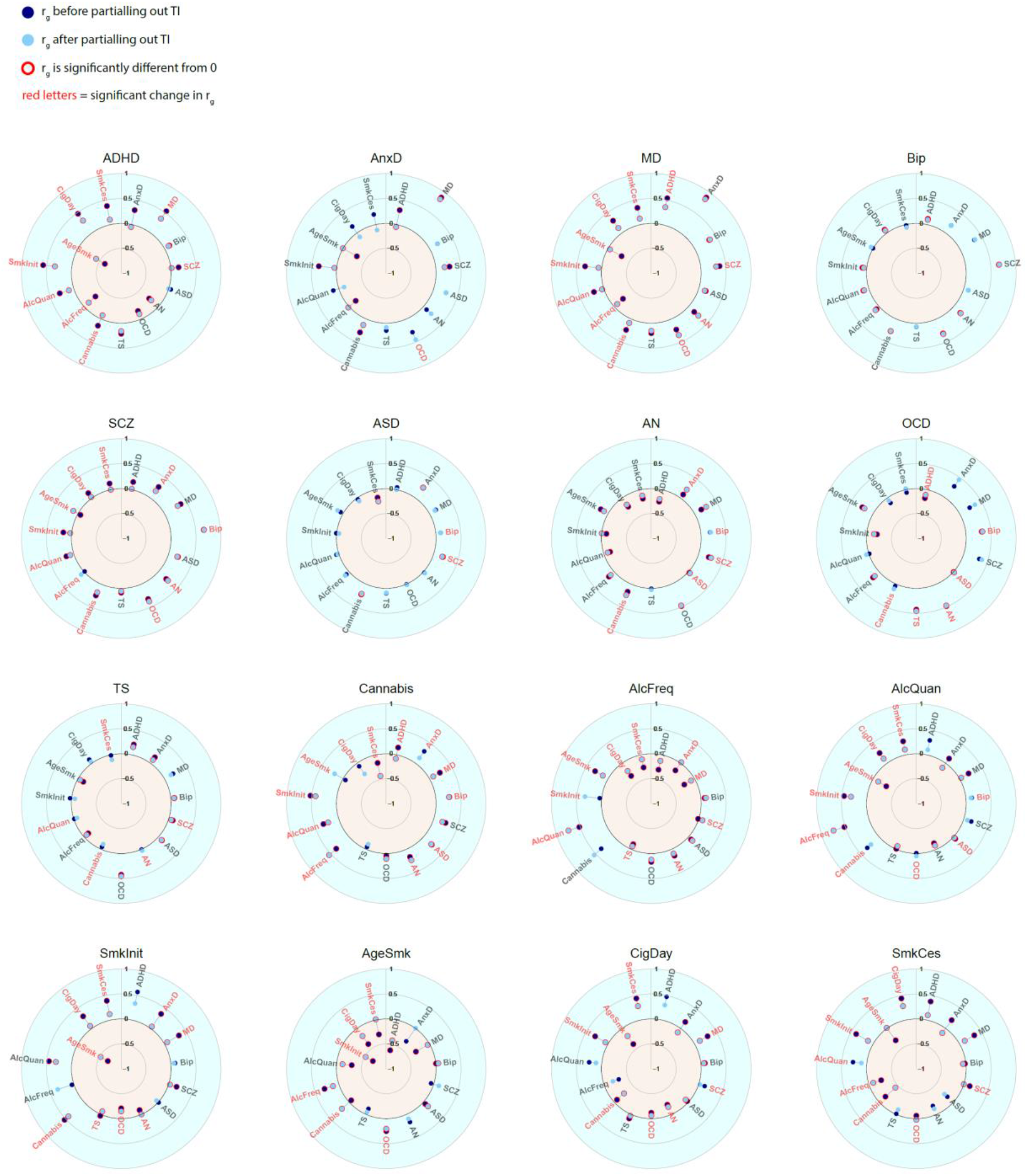
Genetic correlations before and after partialling out Townsend index (TI). Significant genetic correlations are indicated with red circles and significant changes in genetic correlations after partialling out TI are indicated in red letters.

**Supplementary Figure 7:**
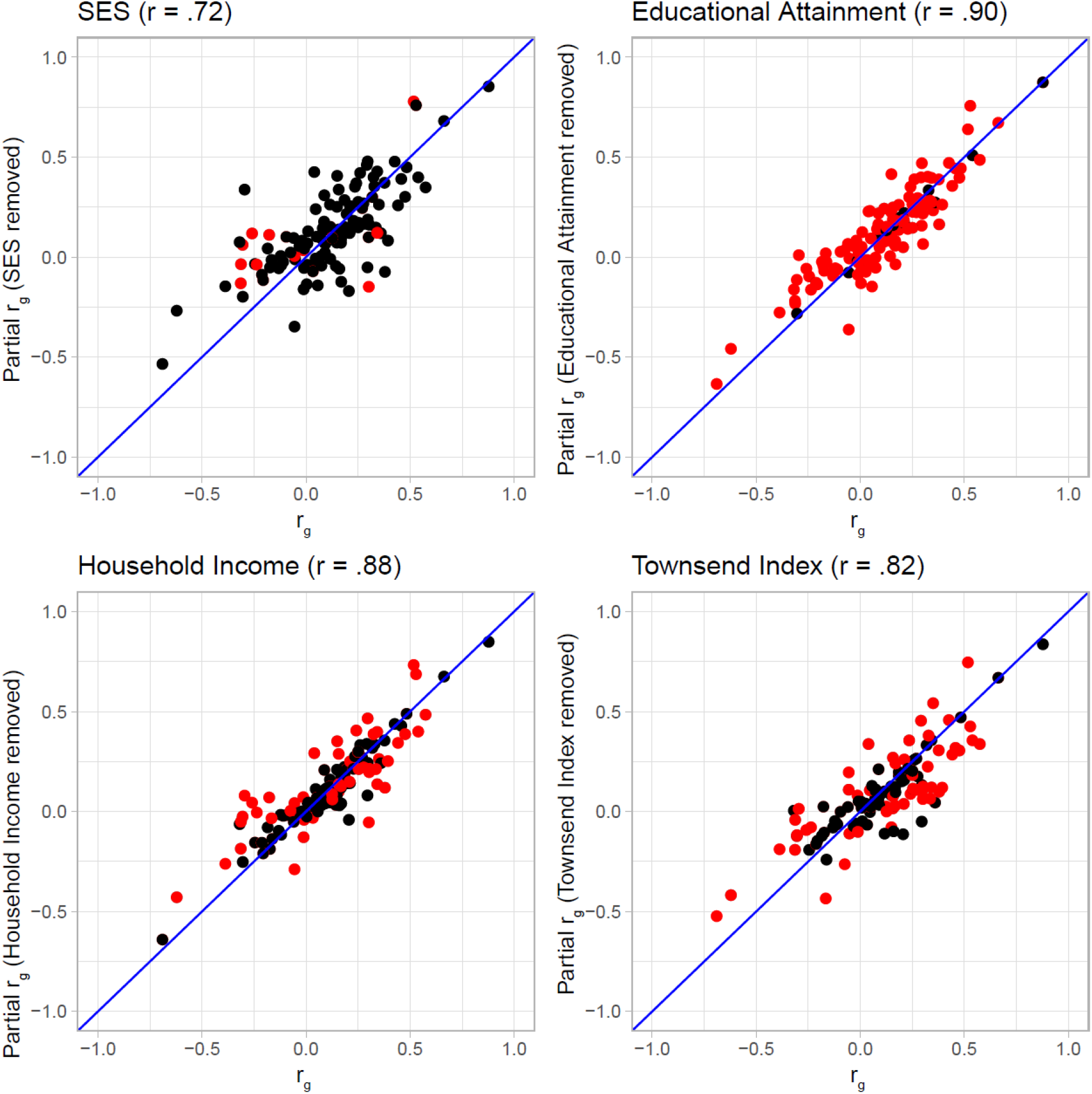
Genetic correlations before (x-axis) and after (y-axis) partialling out SES. Each dot represents one of the mental health or substance use traits. Significant changes in genetic correlations after partialling out SES are indicated as red dots. The four correlations on top of the Figures are the Pearson correlations between the genetic correlations before and after partialling out the SES factors.

**Supplementary Figure 8:**
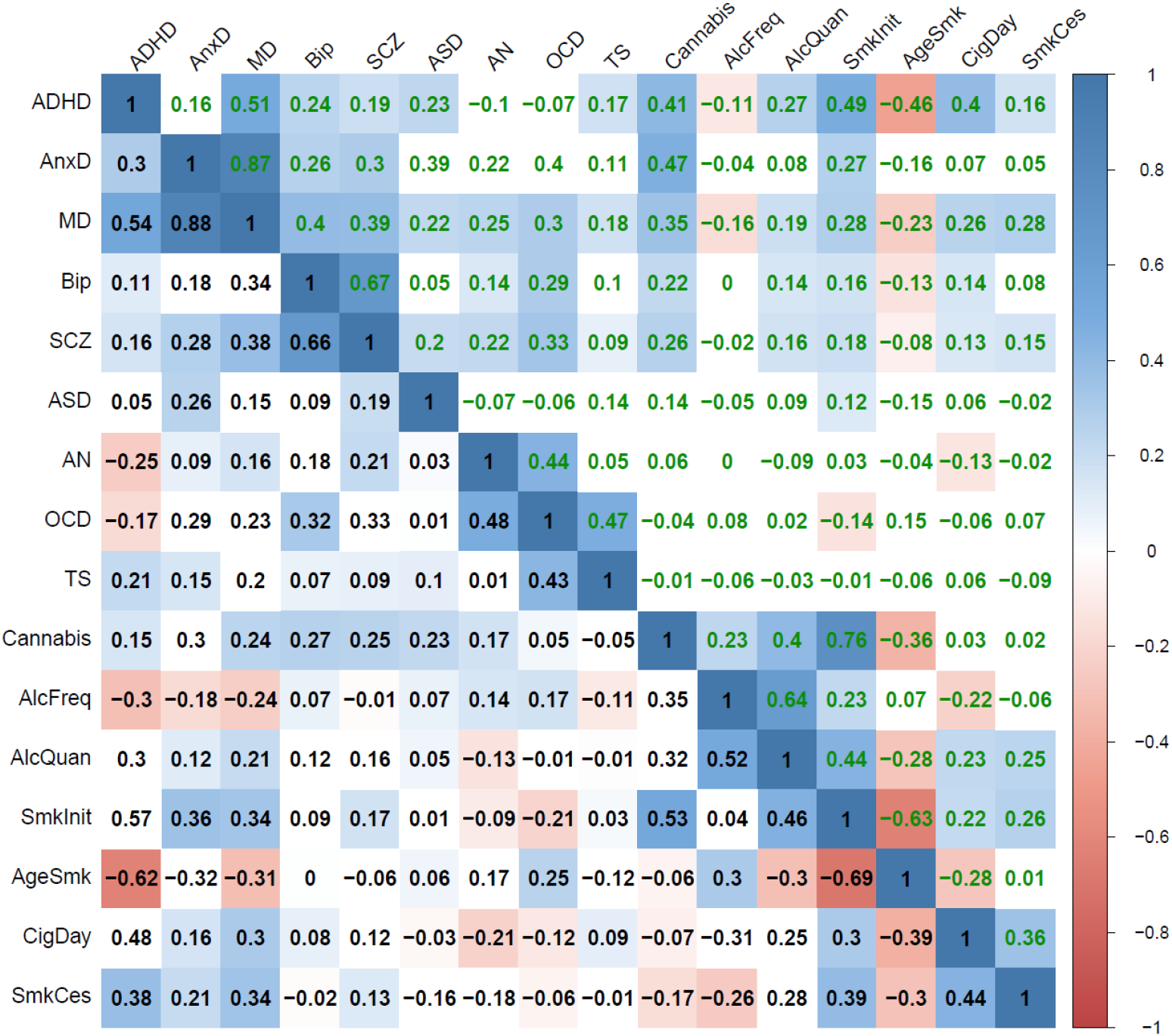
The genetic correlations before (lower diagonal in black type) and after (upper diagonal in green type) partialling out genetic variance of educational attainments. Coloured squares indicate significant genetic correlations (FDR corrected, see methods).

**Supplementary Figure 9:**
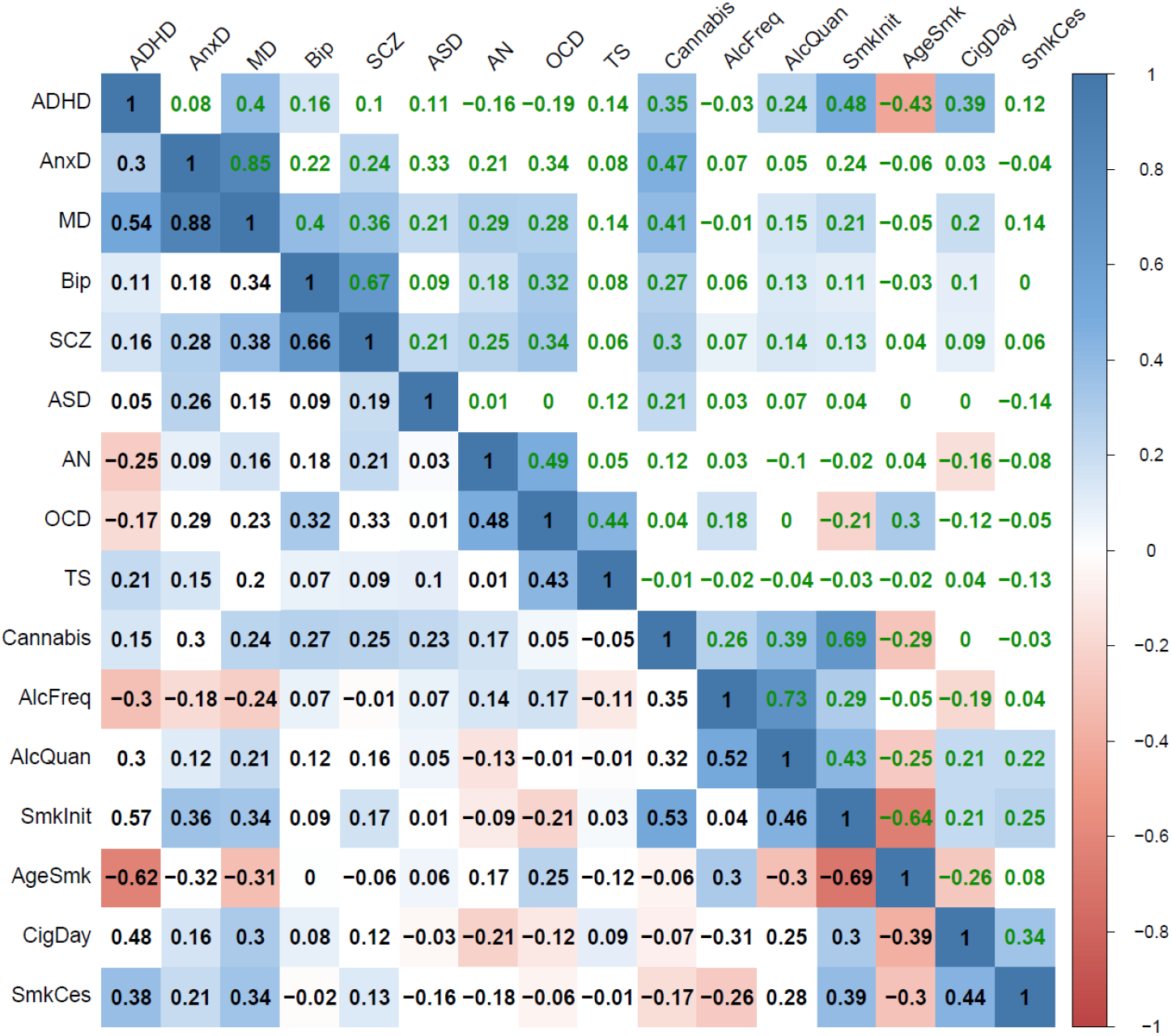
The genetic correlations before (lower diagonal in black type) and after (upper diagonal in green type) partialling out genetic variance of household income. Coloured squares indicate significant genetic correlations (FDR corrected, see methods).

**Supplementary Figure 10:**
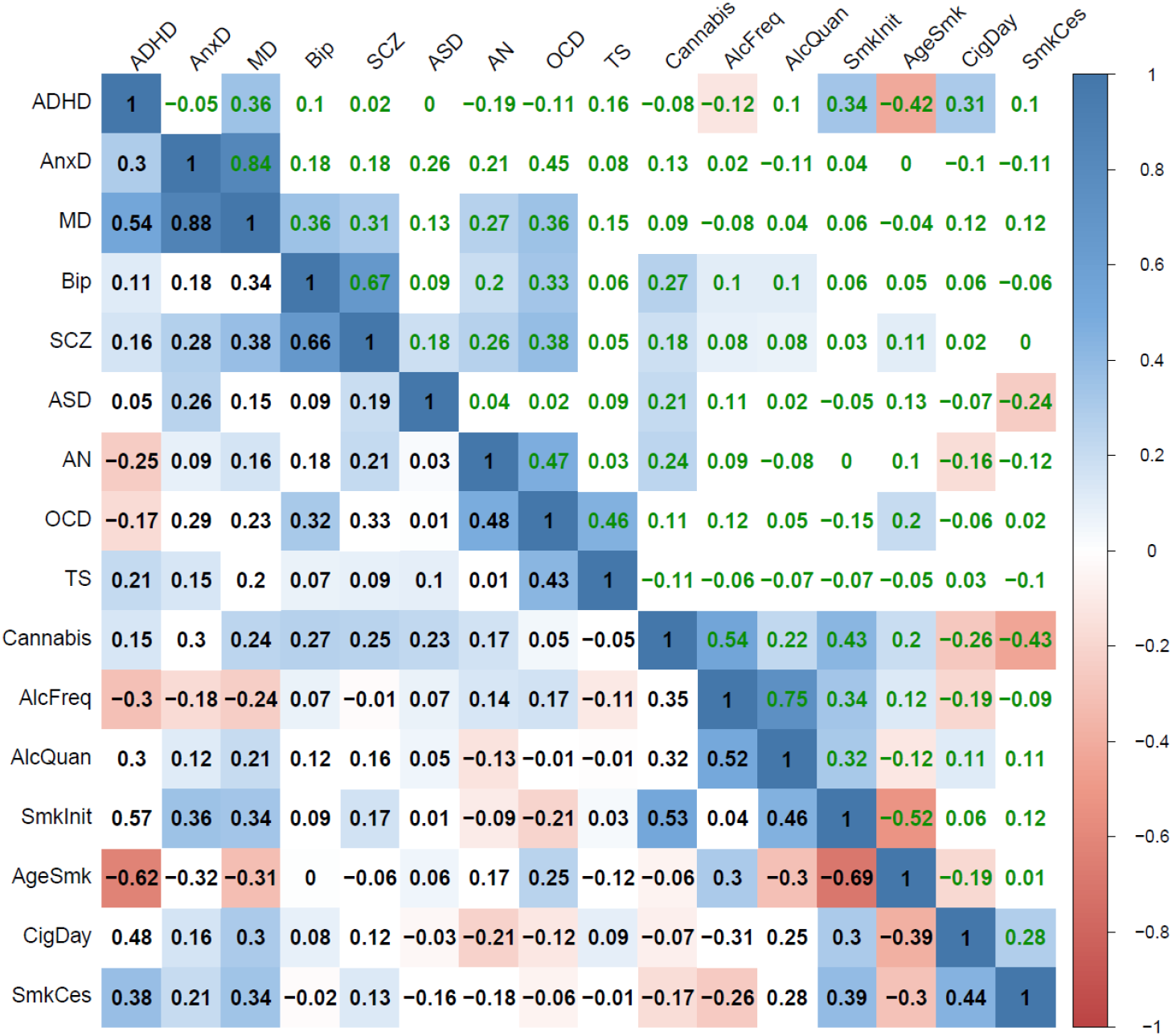
The genetic correlations before (lower diagonal in black type) and after (upper diagonal in green type) partialling out genetic variance of the Townsend index. Coloured squares indicate significant genetic correlations (FDR corrected, see methods).

## Notes

### Competing Interest Statement

The authors have declared no competing interest.

